# RELATIONSHIP OF LIVER ENZYME LEVELS WITH THE CLINICAL COURSE OF COVID-19

**DOI:** 10.1101/2021.03.29.21254559

**Authors:** Ulku İnce, Harun Tolga Duran

## Abstract

**Introduction:** Covid 19 infection, which can affect many systems in the human body, can cause organ dysfunction. High liver serum enzymes can be found in covid-19 patients, and many factors cause this stop. Patients with high levels of liver enzymes that require invasive mechanical ventilation during their follow-up were examined, and it was aimed to determine whether it was among the predictive indicators of mortality.

**MATERIAL AND METHODS:** Patients infected with covid 19 who were hospitalized in the intensive care unit between March 30 and December 1, 2020 according to the criteria of hospitalization in the intensive care unit, clinical trials such as age, gender, length of stay, additional diseases, liver enzyme levels and whether invasive mechanical ventilation is required their characteristics were recorded and analyzed retrospectively and compared.

**RESULTS:** Data were collected from 111 patients whose liver enzyme levels were measured from 131 patients included in the study. It was found that aspartate transaminase, alanine aminotransferase, and gamma-glutamyl transferase levels were statistically higher in the invasive mechanical ventilation group compared to the patients who did not undergo invasive mechanical ventilation.

**CONCLUSION:** Alanine aminotransferase, aspartate aminotransferase, and gamma-glutamyl transferase levels were statistically higher in COVID19-infected patients who were treated in intensive care and undergoing invasive mechanical ventilation. These enzymes are easily accessible and are shown among predictive values in mortality.

## Introduction

Covid 19 infection has been spreading globally and threatening human health since the day it emerged. This epidemic, which emerged in China, spread throughout the world in a short time and affected millions of people. [1] Covid 19 infection infects many systems in the human body and causes organ dysfunction. It causes respiratory distress, especially with pneumonia, and affects the circulatory, hepatic, renal, and hematological systems [2–3]. High liver tests can be found in covid 19 diseases, and the cause is unclear. It is thought that it may develop due to direct pathogenic effects of the Sars Cov-2 virus, side effects of drugs used in moderate and severe covid 19 patients, systemic inflammatory response, or hypoxia. However, there is uncertainty as to whether this elevation in liver enzymes increases the severity of the disease [4]. Some previous studies on sars coronavirus have shown that liver enzymes may be elevated in patients infected with sars [5]. In another study, it was thought that liver enzyme levels such as aspartate aminotranferase (AST) and alanin aminotransferase (ALT) were elevated in patients with covid 19, and this infection could lead to liver damage [6]. Our aim is to examine the laboratory results of patients infected with covid19 in our hospital’s anesthesia intensive care units. To determine whether liver enzymes are among the predictive factors effective in showing the severity of the disease and mortality and to contribute to the literature on this subject. For this purpose, we grouped and examined patients according to their status of receivinginvasive mechanical ventilation (IMV) support, considering that the need for IMV support indicates that the disease is more severe.

## MATERIAL AND METHOD

After the ethics committee approval was obtained between 30 March and 1 December, a total of 131 patients hospitalized in anesthesia intensive care units of our hospital due to covid19 were retrospectively reviewed. Patients with chronic liver disease and lack of data were excluded, and a total of 111 patients were included in the study. After obtaining ethics committee approval for this observational retrospective study, we reviewed the medical records of patients who were followed up and treated in anesthesia intensive care units due to COVID-19. After the patients were taken to the intensive care unit, clinical and biochemical data were collected retrospectively from medical records. The patients who did not receive IMV support were classified as Group 1, and the patients who were administered Group 2.

### Inclusion criteria

A radiologist confirmed patients aged 18 years and over who admitted to our hospital with complaints such as fever, cough, and shortness of breath, and after the diagnostic imaging findings of COVID-19 infection, the diagnosis of covid-19 was supported by the detection of nucleic acid in the respiratory tract by a polymerase chain reaction and met the criteria for admission to intensive care.

### Exclusion criteria from the study

Patients with chronic heart disease, chronic alcohol use, and lack of data were excluded from the study.

### Criteria for admission to intensive care

Respiratory rate above 20 and oxygen saturation of 90 and below despite 100% oxygen support of 5 lt / min with a reservoir oxygen mask or invasive mechanical ventilation support was applied to emergency departments with respiratory distress.

### Invasive mechanical ventilation application criteria

In spite of HFNO or CPAP support, he was taken to invasive mechanical ventilation support in cases with oxygen saturation below 90%, respiratory rate above 20 20, and deterioration of the patient’s hemodynamic findings.

## RESULTS

There were 34 patients in group 1 and 77 patients in group 2. 15 (%) of the patients in Group 1 and 28 of the patients in Group 2 were women. The mean age of the patients in Group 1 was 67 ± 10, while the mean age of the patients in Group 2 was 69 ± 15 (p: 0.00) (Table 1).

**Table 1:**
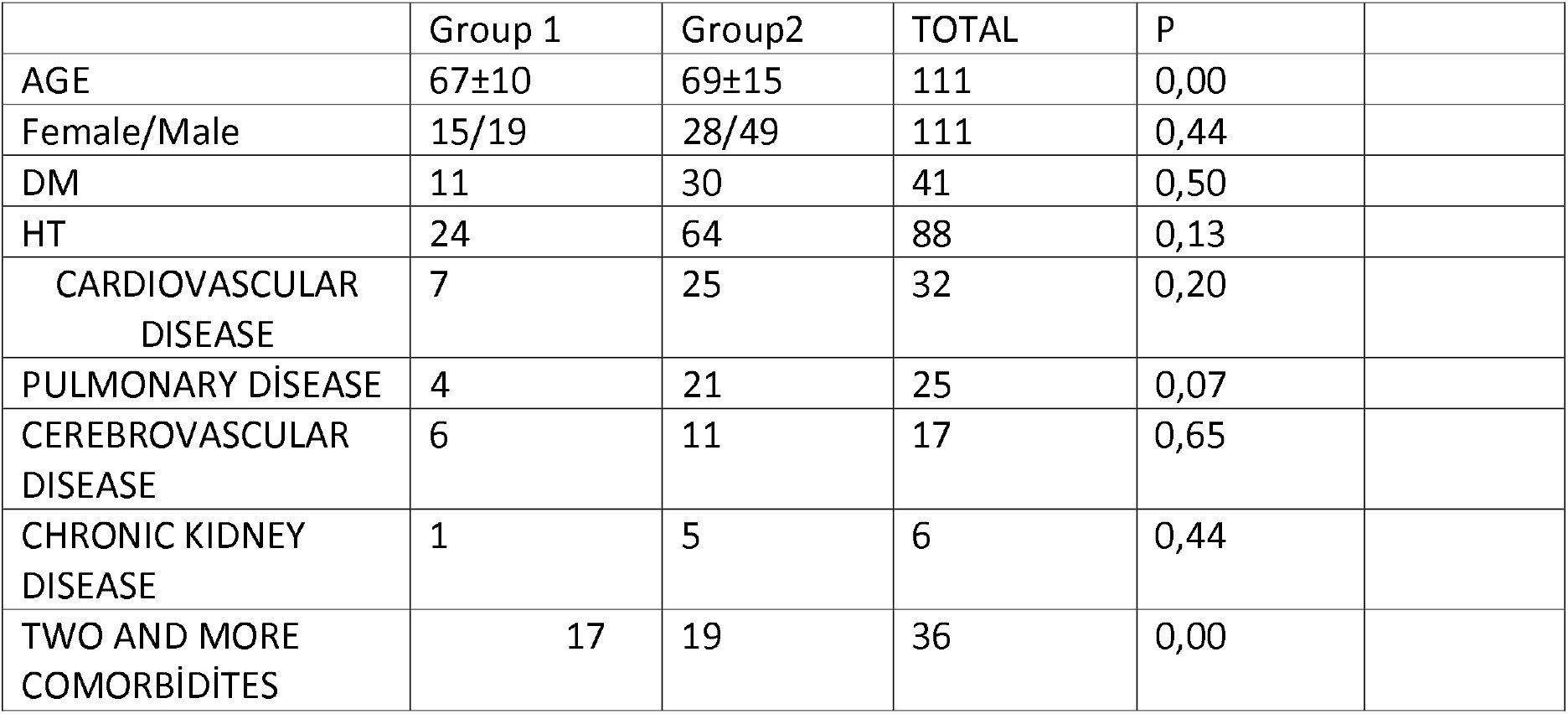
Demographic data and clinical characteristics of the patients.

It was observed that 5 of 77 patients in Group 2 were admitted to the intensive care unit while under invasive mechanical ventilation support. It was observed that the other 72 patients received invasive mechanical ventilation support during their follow-up in the intensive care unit. While all 34 patients in Group 1 were discharged from the intensive care unit, only 4 of the errors in Group 2 were discharged from the intensive care unit (p: 0.00).

ALT, AST, GGT and LDH were significantly higher in Group 2 than in Group 1. Respectively; (p: 0.000), (p: 0.005), (p: 0.006), (p: 0.003). (Table 2).

**Table 2:** Comparison of the laboratory results of the groups

|  | GRUP 1 |  | GRUP2 |  |  |
| --- | --- | --- | --- | --- | --- |
|  | Mean±SD | N | Mean±SD | N | P |
| **Biochemical characteristic** |  |  |  |  |  |
| Albumin g/dl | 3,2± 0,3 | 34 | 3,1± 0,2 | 77 | 0,461 |
| ALT IU/l | 30±9 | 34 | 49±29 | 77 | 0,000 |
| AST IU/l | 22±7 | 34 | 32±16 | 77 | 0,005 |
| GGT IU/l | 29±15 | 28 | 76±80 | 56 | 0,006 |
| ALP IU/l | 62±18 | 20 | 96±49 | 41 | 0,512 |
| D Bil mg/dl | 0,2±0,1 | 33 | 0,2±0,2 | 73 | 0,065 |
| İ bil mg/dl | 0,2±0,2 | 34 | 0,2±0,1 | 73 | 0,330 |
| Glukoz mg/dl | 173±78 | 34 | 185±108 | 77 | 0,218 |
| Bun mg/dl | 67±84 | 34 | 58±47 | 76 | 0,004 |
| Kreatinin mg/dl | 1,1±0,4 | 34 | 1,3±1,8 | 77 | 0,004 |
| T Protein mg/dl | 6±0,6 | 29 | 8±0,1 | 62 | 0,053 |
| LDH IU/l | 338±154 | 30 | 408±187 | 60 | 0,003 |
| Na mmol/l | 136±4 | 34 | 138±3 | 77 | 0,131 |
| Potasium mmol/l | 4,2±0,5 | 33 | 4,2±0,7 | 77 | 0,588 |
| Cl mmol/l | 94±25 | 34 | 102±5 | 77 | 0,279 |
| Ca mg/dl | 8,3±0,5 | 34 | 8,1±0,8 | 77 | 0,033 |
| P mg/dl | 3,0±0,9 | 25 | 2,9±1,1 | 51 | 0,658 |
| Mg mg/dl | 1,9±0,2 | 31 | 2,0±0,2 | 74 | 0,065 |
| **Hematological assessment** |  |  |  |  |  |
| leukocyte cells/mm<sup>3</sup> | 11,4±5,2 | 34 | 10,5±5,0 | 77 | 0,409 |
| Neutrophil cells/mm<sup>3</sup> | 9,9±4,9 | 34 | 9,6±3,9 | 77 | 0,219 |
| Lymphocyte cells/mm<sup>3</sup> | 0,8±0,3 | 34 | 0,7±0,4 | 77 | 0,880 |
| Hgb gr/dl | 11,5±1,4 | 34 | 11,9±1,7 | 77 | 0,012 |
| Rbc fl | 3,9±0,6 | 34 | 4,1±0,5 | 77 | 0,850 |
| Plt cells/μl | 287±130 | 34 | 202±82 | 77 | 0,119 |
| Mpv fl | 10,1±0,8 | 34 | 10,4±1,6 | 77 | 0,015 |
| **Coagulation tests and inflammatory profile** |  |  |  |  |  |
| Inr | 1,1±0,1 | 34 | 1,2±0,3 | 77 | 0,041 |
| Fibrinojen mg/dl | 598±161 | 33 | 660±134 | 75 | 0,010 |
| D-Dimer ng/ml | 1913±1869 | 34 | 1805±2201 | 77 | 0,271 |
| CRP mg/l | 79±60 | 34 | 128±78 | 77 | 0,008 |
| Ferritin ng/ml | 656±684 | 34 | 714±435 | 77 | 0,036 |
| Procalcitonin ng/ml | 0,2±0,4 | 32 | 4,3±16 | 73 | 0,001 |
| İGA ng/ml | 2,6±1,0 | 31 | 2,5±1,1 | 73 | 0,009 |
| IL 6 ng/ml | 32±39 | 32 | 243±360 | 74 | 0,000 |

A positive correlation was found between ALT, AST, GGT and LDH levels and IMV support. Respectively; (p: 0.043, r: 0.193), (p: 0.047, r: 0.189), (p: 0.043, r: 0.222) (p: 0.010, r: 0.272), (Table 3).

**Table 3:**
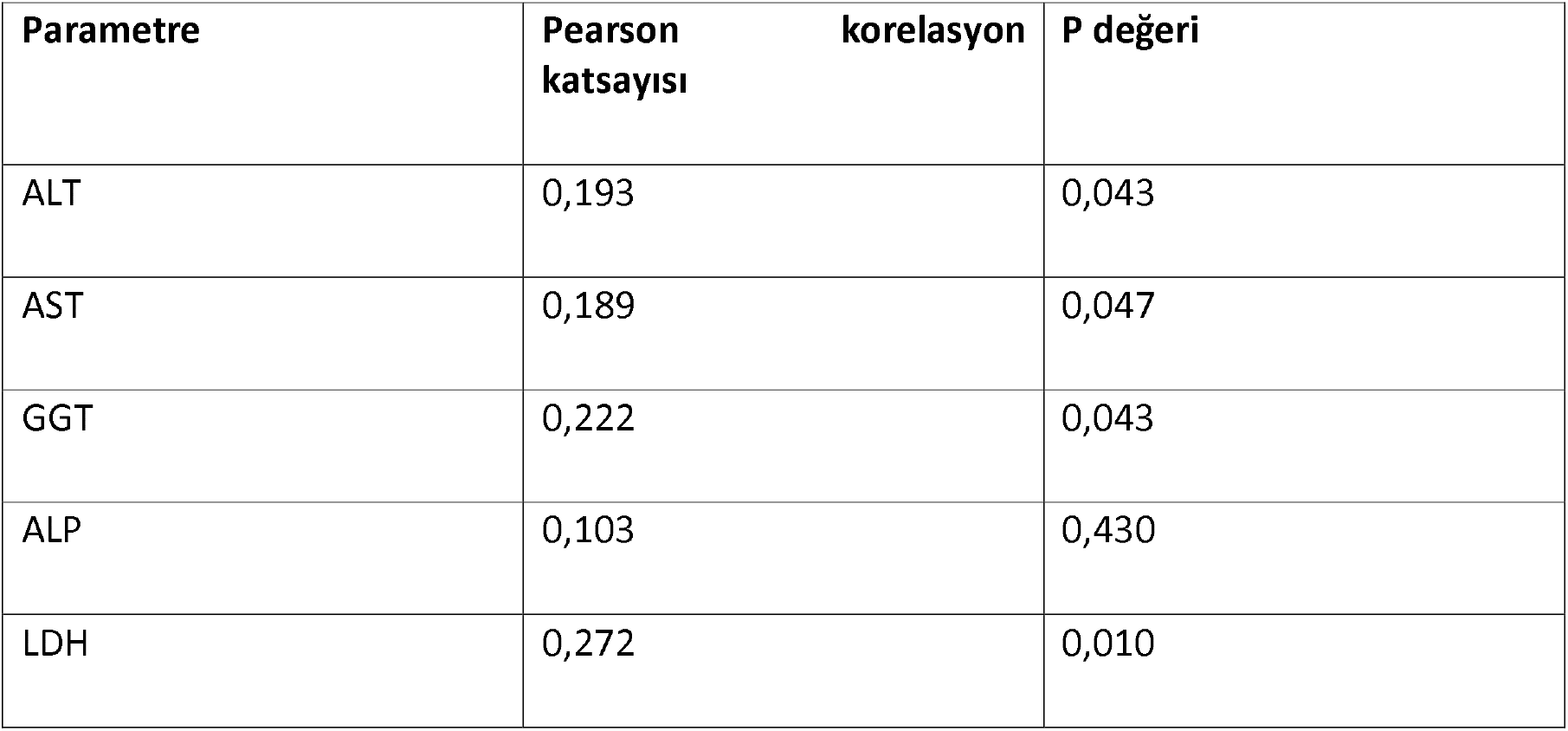
Correlation coefficient and p value between liver enzymes and IMV.

Creatinine (p: 0.042, r: 193), sodium (p: 0.40, r: 195), chlorine (p: 0.44, r: 191), fibrinogen (p: 0.10, r: 247), A positive correlation was found between CRP (p: 0.006 r: 257), age (p: 0.10 r: 243) and IMV support. There was no correlation between other parameters and IMV support.

## Discussion

Due to the constantly increasing number of cases and deaths, it is important to understand the factors associated with mortality from covid 19 infection and the prognosis of this disease. Factors associated with mortality, especially in critical covid 19 patients hospitalized in intensive care units, include age, gender, and the presence of comorbid diseases [7]. In their study, guan et al. showed an increase in the need for invasive mechanical ventilation (IMV). In our study, when gender and diabetes mellitus, hypertension, chronic cardiological neurological, kidney, and lung disease were compared between the groups, there was no statistically significant difference. However, age and the presence of two or more diseases in group2 were statistically higher than in group1.

Covid 19 diseases are associated with elevated transaminases, d dimer, procalcitonin, and some laboratory findings such as troponin [8]. Some previous studies on sars coronavirus have shown that 34% of patients infected with sars may have elevated liver enzymes. N protein and RNA polymerase gene fragments of the SARS virus were detected in hepatocytes in these patients. [34]. Covid 19 infection is thought to cause cellular damage and systemic inflammatory response by binding to ACE 2 receptors in the liver [9]. This binding is made through S proteins on the surface of the virus. It is thought that S2 protein is responsible for fusion while attaching to cells with S1 protein [10] that elevated liver enzyme levels such as AST and ALT can be seen in patients with covid 19, and the appearance of microvesicular steatosis and mild lobularity in the liver in autopsies of some covid 19 patients suggests that liver damage may be caused by this infection [6]. It has been shown that Sars Cov-2 infection is a disease that causes coagulopathy and thrombosis and can cause tissue edema and damage due to diffuse coagulation. Hepatic ischemic perfusion injury is thought to cause inflammation and cell damage by activating Kupfer cells, neutrophils, and platelets. [1-11]. Tijera et al. In their study, it was observed that significant increases in liver enzymes play a very important role in the more severe course of COVID-19 [12]. Similarly, some studies have reported higher mortality in COVID-19 patients with elevated liver enzymes [13-14-15-16-17]. The mortality rate was 88-97% in covid 19 patients who received IMV support [18-19]

In our study, serum liver enzyme levels were found to be statistically higher in GROUP 2. In addition, a positive correlation was found between AST, ALT, LDH and GGT, and IMV support. This situation can be accepted as an early predictive indicator of mortality and the need for mechanical ventilation in patients with high transaminase levels.

It is thought that direct pathogenic effects of the virus, side effects of drugs used in critically ill patients, chronic hypoxia, and systemic inflammatory response may play a role as the cause of elevated serum transaminase levels in patients with Covid 19 [4].

All of the patients included in our study were critical covid 19 patients who were initiated antiviral therapy and were taken to intensive care due to respiratory distress and hypoxia. The antiviral and antibiotic drug groups used and hypoxia may have caused elevated liver enzyme levels.

CRP, Ferritin, IL-6, and procalcitonin levels are also parameters considered as indicators of mortality in Covid 19 patients [20]. In our study, inflammatory cytokines and procalcitonin levels such as CRP procalcitonin and IL 6 were also found to be statistically higher in the group patients who needed IMV support compared to the patients who did not, and this supports that systemic inflammatory response may be more severe.

However, the measurement of transaminases is more practical and applicable; It is more useful alongside other parameters that are limited in use.

## RESULT

There are many factors related to morality in critical covid 19 patients. During the Kovid 19 pandemic, the need for MV and intensive care beds has increased all over the world. It is important to predict the severity of the disease in advance in terms of early preparation and planning of treatment by anticipating the need for mv and intensive care beds. Measurement of liver enzymes is an easy and inexpensive method that can be applied in many centers. In our study, we found that the disease was more severe in patients with high kc enzymes. Elevation of liver enzymes may be one of the predictive factors in showing the severity of covid 19 diseases. More studies are needed.

## Data Availability

Articles cited are available

